# Quality of Life in elderly ICU survivors: A Rapid Systematic Review and Meta-Analysis of Cohort Studies

**DOI:** 10.1101/2020.08.25.20181776

**Authors:** Kevin Ariyo, Sergio Canestrini, Anthony S. David, Alex Ruck Keene, Gareth S. Owen

## Abstract

**BACKGROUND:** The influence of age upon intensive care unit (ICU) decision-making is complex and it is unclear if it is based on expected subjective or objective patient outcomes. To address recent concerns over age-based ICU decision-making we explored patient-assessed quality of life (QoL) in ICU survivors.

**METHODS:** We conducted a rapid database search of cohort studies published between January 2000 to April 2020, of elderly patients admitted to ICUs. We extracted data on self-reported QoL (EQ-5D composite score), study characteristics and demographic and clinical variables. Using a random-effects model, we then compared QoL scores at follow-up to scores either before admission, age-matched population controls or younger ICU survivors. Finally, we conducted follow-up quantitative analyses to explore potential moderators of these effects, and a qualitative synthesis of QoL subscores. A study protocol was registered prospectively on PROSPERO, ID: CRD42020181181.

**FINDINGS:** Our database search found 2536 studies and from these we reviewed 376 potentially relevant full texts. 21 of these studies met the inclusion criteria for qualitative synthesis and 18 were also included in the meta-analysis (N= 2090 elderly adults). The follow-up periods ranged between 3-100 months. There was no significant difference in the elderly’s QoL scores between one month before ICU and follow-up, or between follow-up and age-matched community controls. QoL in elderly ICU survivors was significantly worse than younger ICU survivors, with a small-to-medium effect size (d= .33 [.10 to .55]). Mortality rates and length of follow up were possible intermediary factors. The qualitative synthesis suggested that any reductions in QoL were primarily due to reductions in physical health, rather than mental health items.

**INTERPRETATION:** Overall, elderly ICU patients did not experience significantly impaired QoL at follow up, compared to before ICU or their healthy peers. Elderly patients who survive ICU can be expected to have slightly worse QoL compared to younger patients, especially in the longterm. The results suggest that the proportionality of age as a determinant of (population level) ICU resource allocation should be kept under close review and that subjective QoL outcomes (not only objective survival data) should inform person-centred decision making in elderly ICU patients.

**DECLARATIONS OF INTEREST:** Alex Ruck Keene is an adviser on the Faculty of Intensive Care Medicine’s Legal and Ethical Policy Unit. We report no other competing interests.

## INTRODUCTION

The influence that age should have upon intensive care decision making has been debated across policy and clinical practice ^1 2^. Age associates (inversely) with the probability of intensive care unit (ICU) survival and length of life after ICU ^3 4^, outcomes generally considered to be relevant to resource allocation ^2^. However age is also a protected characteristic in several jurisdictions, and in England and Wales, resource allocation based on age must be a “proportionate means of achieving a legitimate aim”, if it is not to be contrary to the Equality Act (2010).

For elderly patients for whom admission to ICU is clinically appropriate, an important part of person-centred decision-making is for them, or their families, to be given information about the likely outcome of admission. Patients may seek to integrate survival and biomedical outcomes with subjective outcomes, including quality of life (QoL). Subjective QoL in elderly ICU survivors has been studied less frequently than these objective measures ^3 5^. This is notable given that subjective QoL (via Quality-Adjusted Life Years, or QALYs) is very influential in clinical resource allocation (e.g. NICE). Person-centred decision making requires consideration of patient experience since physician-rated quality of life is not always well correlated with patient-rated quality of life.

We considered a rapid review to be urgent because age is a strong risk factor for severe COVID-19 infection ^6^ and severe COVID-19 has placed considerable pressure on ICU resource allocation. ^7^ and is likely to do so in the future. Additionally, some have expressed concerns that elderly adults may be disproportionately less likely to receive ICU ^1 2 8-10^. It is therefore important older persons’ subjective outcomes are better understood.

We conducted a meta-analysis on patient reported QoL in elderly adults undergoing ICU. Following a systematic review, we addressed three questions:

1. At follow up, do elderly ICU survivors have better/worse QoL compared to their scores before ICU?
2. At follow up, do elderly ICU survivors have better/worse QoL than age-matched community controls?
3. At follow up, do elderly ICU survivors have better/worse QoL than ICU survivors aged under 65?

Determining the effect of illness and ICU on QoL is complicated because QoL is itself influenced by many variables ^11^ and some are non-clinical. These influences are too complex to resolve completely, but where possible, we sought to model relevant variables (illness severity, ICU length of stay and mortality rate) as predictors of QoL in elderly ICU survivors at follow up, compared to controls.

## METHODS

### SEARCH STRATEGY

We searched for English-language journal articles, published between January 2000 and April 2020. Six online bibliographic databases were used: CENTRAL, CINAHL, Cochrane Library, EMBASE, MEDLINE and PsycINFO. Our search followed pre-published PROSPERO protocol (ID: CRD42020181181).

The search terms focused on intensive care, elderly adults and QoL. We supplemented this with a forward citations and reference list search based on the eligible articles as well as consultation with experts.

### SELECTION CRITERIA

We undertook study selection using EndNote X9 using a standardised CRIB sheet. At the title and abstract level, we identified potentially eligible studies that took place in an ICU and referred to either QoL life or elderly adults. Full texts were eligible if a) all participants underwent ICU; b) there were at least 20 elderly patients and controls; c) scores from a validated QoL scale were reported, for a group aged at least 60+, with at least 3 months follow up review; d) the follow up QoL scores were derived from the patient, rather than a professional; and e) the study reported QoL scores from the same scale for either the same patients before the ICU admission, age-matched community controls or ICU survivors aged under 65.

We considered whether to include studies that focused only on cardio or neuro-surgical patients, given the effects of the diagnostic heterogeneity that characterises the reference population of the studies included in our review (general ICU patients with various conditions). However, none of these studies met the other inclusion criteria.

K.A led the study selection at all stages and a post-doctoral research assistant conducted reliability checks for 50% of full text articles. We found nearly perfect inter-rater agreement, as measured by Cohen’s kappa (k= .86) ^12^. Queries were resolved by G.O.

### DATA EXTRACTION

One reviewer (K.A) extracted relevant data from all eligible studies, recording this on a standardised spreadsheet. M.K. independently extracted data from 10% of eligible studies, to evaluate consistency. The primary outcome was the QoL composite scores. Secondary variables included demographics, QoL subscale scores, mortality (from ICU to follow up), illness severity (APACHE-II or SAPS-II), length of ICU stay, length of hospital stay, and average follow up time. When one dataset was used for multiple studies, we included the study with the clearest data reporting.

To ensure consistency, we included only composite scores from the EuroQoL health related quality of life instrument (EQ-5D) within the meta-analysis. Where possible, we also converted the eight SF-36 subscales to an EQ-5D index score, using an established mapping algorithm. ^13^. The remaining studies were included within the qualitative synthesis only.

### DATA ANALYSIS

We explored the effect of age on EQ-5D composite scores using random effects meta-analyses. KA conducted the analysis using R Statistics. We used the Restricted Maximum Likelihood (REML) method to calculate the effect sizes (Cohen’s d), which were weighted by the inverse of the sampling variance: meaning that studies with higher variance contributed less to the summary effect size. We interpreted these effect sizes using conventional criteria as a guide (0.2 = small; 0.5 = medium; 0.8 = large) ^14^. We then conducted sensitivity analyses for each meta-analysis to assess risk of bias at the study level, including heterogeneity (e.g. I^2^ statistic), influential studies (e.g. Cook’s distance), and publication bias (funnel plots and Egger’s test).

To investigate the remaining heterogeneity, we then conducted two secondary analyses: a moderator analysis to explore variation within a specific predictor, and a random-effects meta-regression to explore relationships between multiple predictors.

We used several strategies to handle missing data. When the study only reported median values and interquartile ranges, we estimated the mean and standard deviation using conventional formulae ^15 16^. When neither the standard deviation nor interquartile range was reported, we estimated the standard deviation using prognostic imputation ^17^. This calculates the average of observed variances to estimate the missing standard deviation values. We excluded studies with missing data if these methods were inapplicable.

One reviewer (K.A) assessed the methodological rigour of the included studies using an 11-item quality checklist (three irrelevant items were excluded) ^18^. The criteria were scored as either 2 (complete fulfilment), 1 (partial fulfilment) or 0 (not fulfilled). We then calculated a total score for each study and rated them as either high quality (17/22 or higher), moderate quality (between 10/22 and 16/22) or low quality (9/22 or lower). Queries were resolved through discussion with G.O and S.C.

For the qualitative synthesis, we defined a set of criteria for each measure to allocate subscores to either ‘mental health’ or ‘physical health’ categories. We then calculated a crude average for subscales within these two categories and weighted them on a scale of 1-100 (0= minimum QoL; 100 = maximum QoL). As this approach is subjective, we present these findings only as a qualitative supplement.

This study follows methodological guidance from PRISMA.

**Figure 1.**
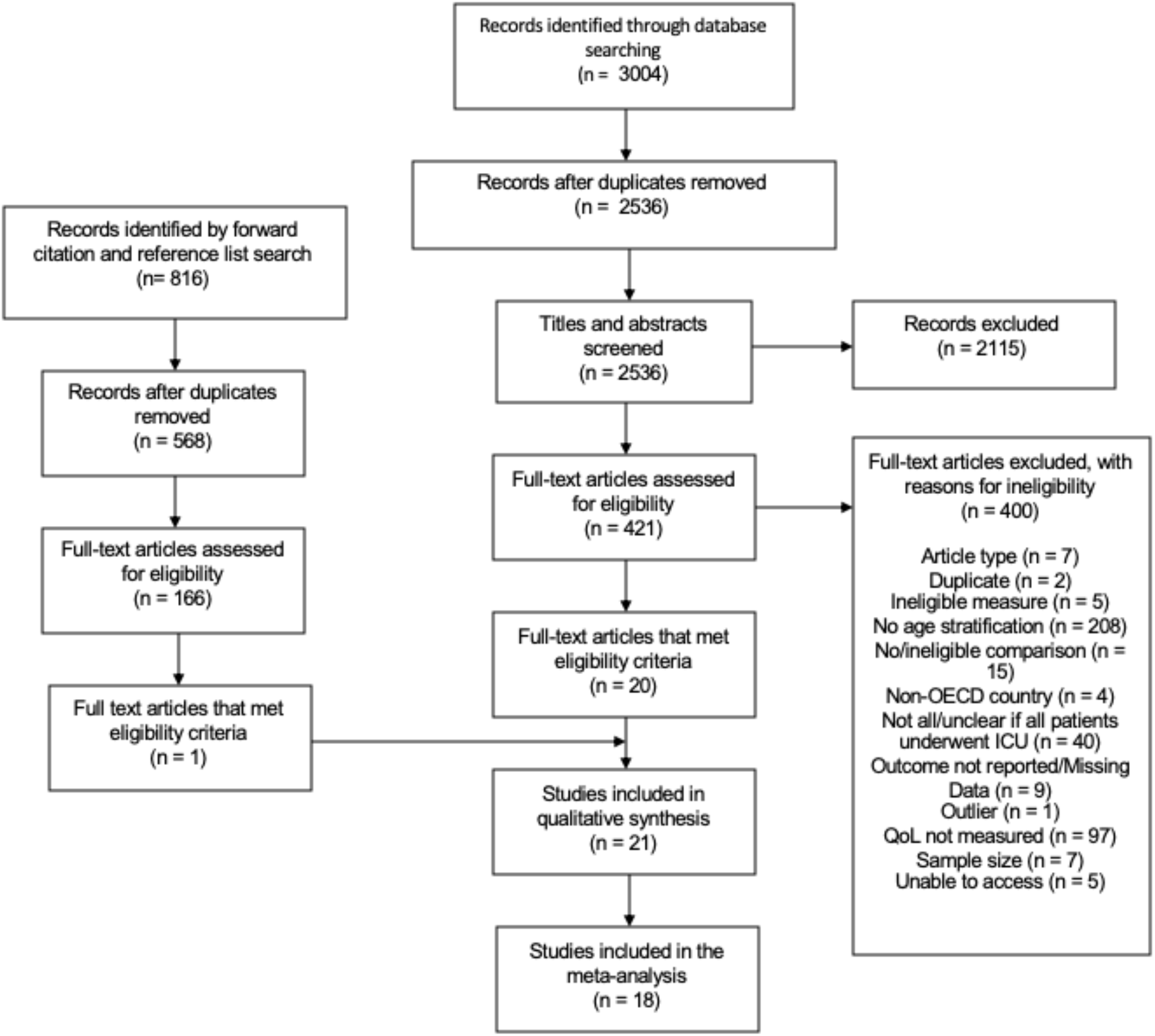
A PRISMA flow diagram that outlines the study selection process.

## RESULTS

### DESCRIPTIVE STATISTICS

After screening duplicates, the database search revealed 2536 records for title and abstract screening. From these, we reviewed 421 potentially relevant full text articles for eligibility.

18 studies met the full criteria and were included in the meta-analysis (N= 2090 elderly ICU survivors) ^19-39^. Eight of these studies reported age characteristics for the elderly patients (M= 78.53, SD= 4.17), while the others reported the minimum age only.

Most of the studies included both medical and surgical ICU patients (fifteen studies). The remaining studies focused on surgical (two studies) or medical (one study) patients only. Three types of outcome were included in the meta-analysis. These results compared QoL at follow up to either pre-ICU scores (five studies), age-matched community controls (nine studies), or younger survivors of ICU (six studies). We provide a full summary in Table 1.

**Table 1.**
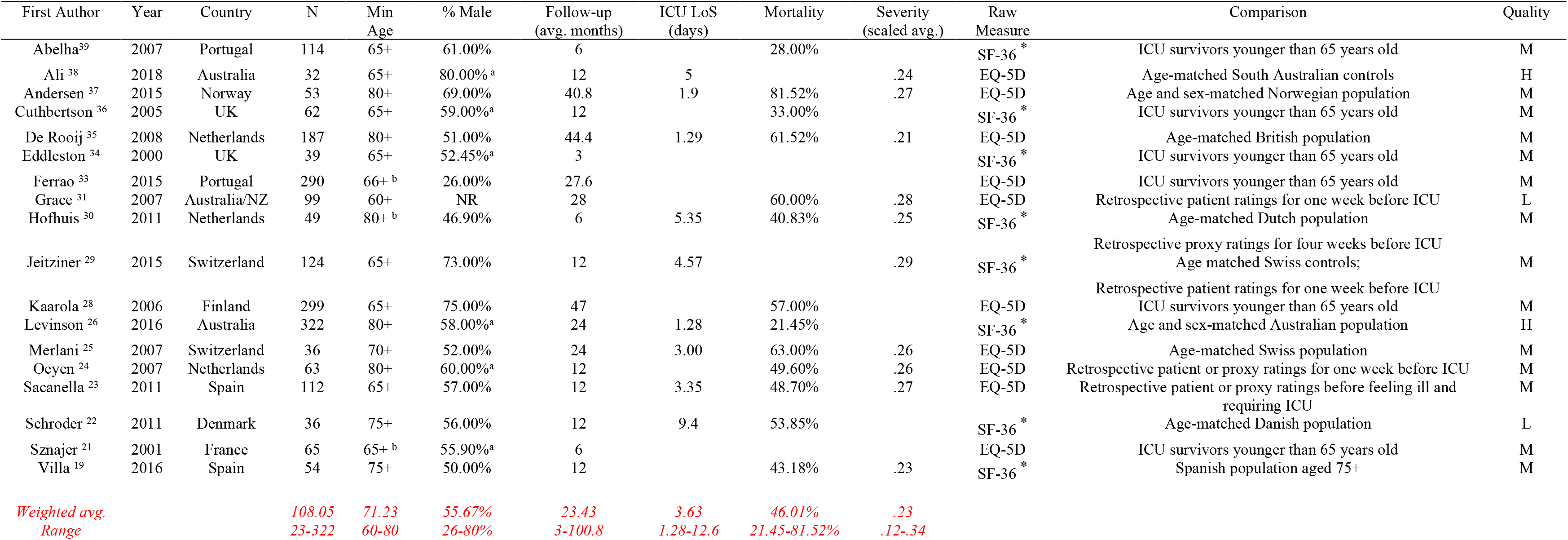
The main characteristics of the studies and the relevant data included in the meta-analyses. ^a^ Reported for study level only, so not included in meta-analysis ^b^ Combined elderly groups * Converted to EQ-5D composite score Abbreviations: ICU (intensive care unit); LoS (length of stay); H = High quality; M= Moderate quality; L= Low quality. See above for measures. Unless specified, we do not report data where it is not representative of at least 66.67% of the included sample.

For the qualitative analysis, four different measurement scales were reported: the EuroQoL EQ-5D health related quality of life instrument (ten studies), the short form medical outcome questionnaire (SF-36; eight studies), the Nottingham health profile (NHP; one study), the quality of life index (QLI; one study) and the World Health Organisation quality of life instruments (WHO-QOL-BREF; one study). SF-36 scores were converted to EQ-5D index scores for the meta-analysis, while the other measures were excluded (see methods).

Table 2 outlines the results of the three meta analyses. There was no significant difference in EQ-5D composite scores between elderly patients before and after ICU (d= -.18, n.s). There was also no significant difference in EQ-5D composite scores between elderly ICU survivors and age-matched community controls (d= -.15, n.s). These results suggest that there were no average differences in QoL between these groups.

**Table 2.**
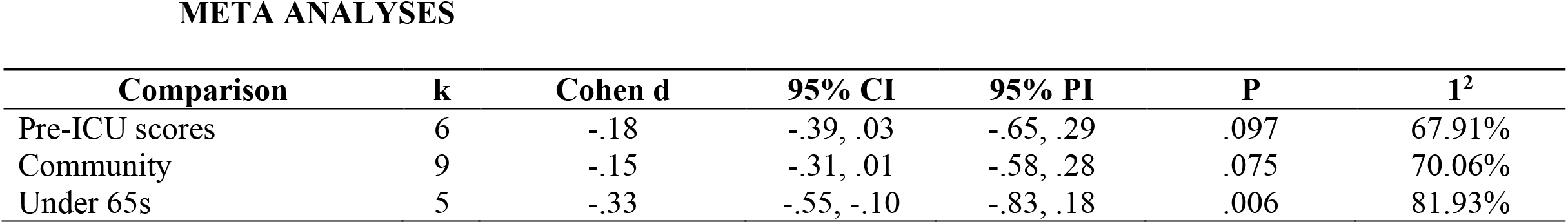
A summary of effect sizes, confidence intervals, prediction intervals, significance and heterogeneity for each meta-analysis (k= number of independent samples, I^2^= between study heterogeneity)

| Comparison | k | Cohen d | 95% CI | 95% PI | P | I<sup>2</sup> |
| --- | --- | --- | --- | --- | --- | --- |
| Pre-ICU scores | 6 | -.18 | -.39, .03 | -.65, .29 | .097 | 67.91% |
| Community | 9 | -.15 | -.31, .01 | -.58, .28 | .075 | 70.06% |
| Under 65s | 5 | -.33 | -.55, -.10 | -.83, .18 | .006 | 81.93% |

Elderly ICU survivors (aged over 65) had significantly lower composite scores on the EQ-5D, compared to younger ICU survivors (aged under 65), with a small-to-medium effect size (d= -.33, p= <.01). This suggests that on average, QoL in elderly ICU survivors is slightly worse than younger ICU survivors.

### SENSITIVITY ANALYSES

We reviewed the impact of influential cases within each analysis. One study was excluded from the community meta-analysis as a substantial outlier and influential result ^40^. If the result had not been excluded, the effect size would have been stronger (d= -1.80 – ie a larger difference in QoL favouring younger controls) but non-significant (p= .27), mainly due to large heterogeneity (I^2^ = 100%). It is unclear why this study reported substantially outlying results, although the reported standard deviations were considerably lower than other studies.

After excluding this, one other study was marginally influential within the community analysis (see Appendix). ^29^. This study was retained as the between study heterogeneity was moderate and excluding the case would have had little impact on the effect size or interpretation. We identified no further outliers according to our criteria.

### SECONDARY ANALYSES

There was moderate-to-large heterogeneity between studies, therefore we explored the role of other variables using post-hoc subgroup analyses and meta-regressions. These results should be interpreted with caution, due to low sample sizes.

Length of follow up significantly predicted greater differences in QoL between elderly ICU survivors and patients aged under 65 (k= 5, p< .0001). This suggests that elderly survivors may have worse QoL in the long term and comparable QoL in the medium term.

Mortality rate significantly predicted greater differences in QoL between elderly ICU survivors and age-matched community controls (k= 7, p= .01). This revealed that elderly patients had worse QoL than controls in studies with high mortality rates, compared to studies with low mortality rates.

Controlling for these variables reduced heterogeneity between studies to 0% in both cases. No model significantly accounted for variance when the outlier ^40^ was included in the community analysis.

Neither severity of illness, year of publication, sex nor minimum age significantly accounted for heterogeneity between the studies, either individually or within a meta-regression (p>.05).

### RISK OF BIAS

We found no evidence of publication bias for the community or pre-ICU meta-analyses, from either funnel plots or Egger’s test (all p> .05). Most studies had a moderate degree of methodological quality (14/18). We had insufficient power to explore the effect of study quality on quantitative outcomes.

### QUALITATIVE SYNTHESIS

To compare different aspects of QoL, we categorised the subscales into either mental or physical health QoL and calculated a scaled average to enable comparisons (see Table 3). 16/21 studies reported the subscales for both conditions. Our estimates suggest that elderly ICU survivors reported higher average scores on mental health items (M= 57.90/100) than physical health items (M= 50.99/100). Trends in physical health scores compared less favourably to age-matched community controls than did mental health scores (mean differences = -5.23 and -1.71, respectively). Trends in physical health scores were also lower in comparison to younger ICU controls (mean difference = -1.40) whereas mental health scores were higher (mean difference = 2.98).

**Table 3.**
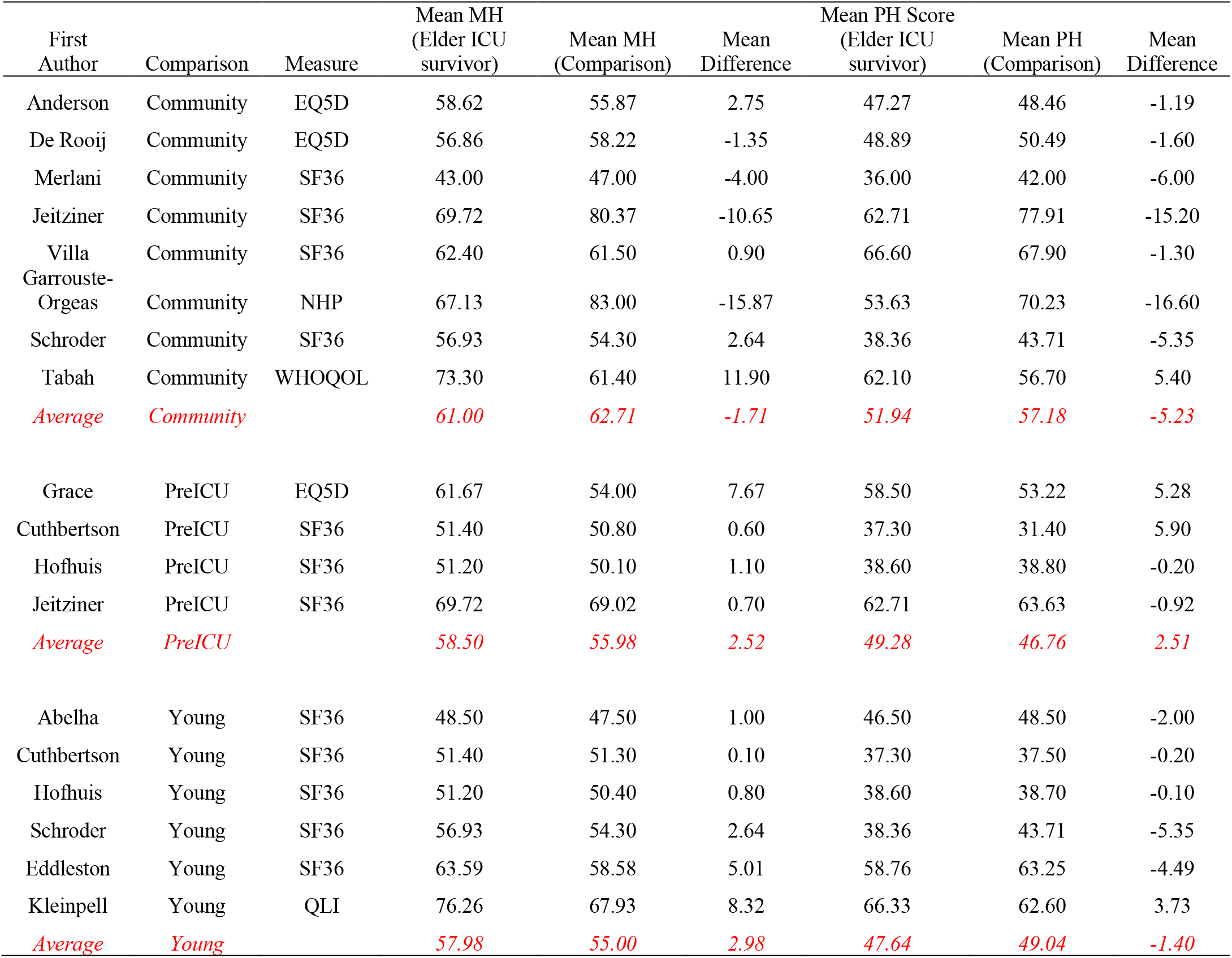
An overview of Quality of Life subscores, by mental health and physical health categories, for elderly ICU survivors and comparison groups. All scores were recalculated on a 0-100 (0 = minimum QoL; 100 = maximum QoL). Abbreviations: MH= Mental Health; PH=Physical Health

## DISCUSSION

This review has systematically evaluated the literature on QoL for elderly ICU survivors in the medium to long term, using EQ-5D composite scores. To our knowledge this is the first meta-analysis to address this issue. We found no evidence of worse QoL after ICU, compared to a period before ICU or compared to healthy community peers. However, elderly patients who survive ICU can be expected to have slightly worse QoL, compared to younger survivors. The wide prediction intervals also suggest that age differences can vary considerably in either direction.

### STRENGTHS IN RELATION TO THE LITERATURE

For the meta-analysis, we identified 2090 elderly ICU survivors within an international sample of 18 cohort studies. We only included recent studies that used validated QoL measures and we rated most studies as having moderate or higher methodological quality. By pooling these samples using rigorous methods, we have been able to overcome several methodological limitations associated with generalising from individual studies, including small samples, choice of analysis and site selection bias. Our sensitivity analyses showed that the remaining heterogeneity was mostly due to conceptually relevant variables. Given the relatively small literature, these methods ensure that valid, transparent results inform policy and clinical practice decisions.

Although contested, previous reviews have generally concluded that age alone is not a suitable determinant of potential benefit from ICU, especially for survivors ^3 5 41 42^. The present study supports these conclusions overall, although the differences compared to younger ICU survivors are still noteworthy. Decisions on whether to admit patients can be extremely difficult for all involved, with seriously ill elderly people overrepresented among the most contentious cases ^43^. These challenges are amplified further when healthcare resources are under pressure, such as during the COVID-19 pandemic.

The age-QoL associations we have found may be explained by intermediary variables. Some research suggests that frailty may best explain age differences in QoL following ICU ^5 44^, and clinical outcome in COVID-19 patients ^45^. Frailty is a more integrative approach to conceptualising ageing, but it was not reported within the eligible studies. We would recommend a meta-analysis of individual patient data to further stratify clinical variables of interest, including frailty, to better predict QoL outcomes.

Health economic analysis of ICU in the elderly based on QALYs may be informative when it comes to resource allocation policies but we have found few such analyses and no explicit polices based on them. They will have to grapple with the controversial notion that everyone is entitled to a ‘normal’ span of health or ‘a fair innings’ ^46 47^. Given the presumption that a sizeable proportion of elderly survivors will enjoy a good QoL it is crucial that holistic, person-centred decision making is not crowded out by survival statistics or anticipatory triage. If triage were to become necessary on the front line we would advise against weighing age too heavily and rather taking more account of frailty after appropriate consultations.

On average, QoL scores gradually decline with age at approximately 0.5 points per year on the CASP-19 (range 0-57) with a modestly accelerated decrease with older age (>85 years) ^4^. It is relevant to consider whether change in QoL in the elderly is primarily due to physical health and mental health components. We were unable to incorporate physical and mental subscores into the meta-analysis due to differences in the levels of data between measures, so we performed a qualitative synthesis. This suggested that for elderly ICU survivors, mental health questionnaire items were relatively unaffected. The small literature on older adults also suggests relatively low rates of anxiety ^48^ and depressive disorders ^49 50^, although potentially high rates of post-traumatic stress. ^51^. Together with previous research, which found that elderly people typically value their psychosocial wellbeing above their physical needs ^52^ our results highlight the importance of caution with assumptions on age as a determinant of poor quality of life following ICU.

### LIMITATIONS

The primary limitation is the small number of eligible studies for each analysis. To maximise the sample, we included some studies with a small amount of missing data and used validated methods to estimate the mean or the standard deviation from the reported statistics. We argue that these approaches are justified as, based on central limit theorem, we expect the larger sample sizes to produce a better estimate of population variance ^53^. For balance, we have also provided a comprehensive overview of our sensitivity analyses to assess risk of bias (see Appendix). These demonstrate that although our decisions reduced bias, most did not change our interpretation of the effects.

Another potential limitation of the meta-analysis is the focus on long-term ICU survivors, as reported mortality rates were as high as 80% at follow up. We argue that a substantial ‘healthy survivor’ effect on QoL is unlikely because survival and QoL have different pathophysiological determinants. We also did not find any evidence of better QoL for elderly patients in studies with high mortality rates. Nevertheless, our results clearly extend only to ICU survivors, rather than prospective ICU patients.

Our results may also be prone to other selection biases. Compared to younger adults, unhealthy elderly adults might be less likely to be admitted to into ICU ^32 43^, to survive ICU treatment (possibly in part due to decisions around lifesaving treatment ^54^) and to survive until follow-up. It was also unclear how many patients had pre-existing cognitive impairments where QoL measurement is more complex, although there was no indication that the proportion was large. As a result, we would caution wider generalisations to all elderly ICU patients. Nonetheless, these results imply that at least a sizeable subgroup of elderly ICU patients will report subjective outcomes that compare well to groups that might be expected to fare better.

We were unable to assess change in quality of life as rigorously as we would have liked. Ideally, we would have analysed differences in QoL change scores between younger and elderly ICU survivors, at multiple time points from before ICU to follow up. The scores for pre-ICU scores were also problematic, as these were determined by retrospective ratings from discharged patients or proxies. This is usual practice, but the reliability of proxies is contested ^55 56^.

Finally, we observed moderate-to-high levels of heterogeneity between studies, which limits the generalisability of the results. We found that much of this variation may have been due to mortality rates and length of time post-discharge, which supports the view that age alone is not a strong predictor of QoL outcome. We also tried to ensure consistency of measurement by using a mapping function between SF-36 scores to EQ-5D scores, which is a common approach within NICE guidelines^13 57^.

## CONCLUSION

Our study reports the first known meta-analysis of quality of life in elderly patients following ICU. We report that on average, elderly survivors of ICU have similar QoL after ICU compared to before and that their QoL is comparable to their community peers. They have slightly worse QoL compared to younger ICU survivors based on physical rather than mental health, but it does not change for the worse following ICU. These findings add rigour to the current literature and should inform debates around population level resource allocation and person-centred intensive care decision making during the current COVID-19 pandemic and after.

## Data Availability

All data is publicly available.

## CORRESPONDENCE

Mr. Kevin Ariyo MSc (PhD candidate), Mental Health and Justice Project, Department of Psychological Medicine, Institute of Psychiatry, Psychology and Neuroscience, King’s College London. 16 De Crespigny Park, Camberwell, London, SE5 8AF. Email.

## AUTHOR STATEMENT

K.A. led at each stage of the project, including drafting the document. G.O. was primary supervisor on the project, jointly formulated the research questions, led on writing the introduction section and made substantial contributions to all aspects of the study. S.C. advised on the initial protocol and provided critical revisions from an intensivist perspective. A.D. and A.R.K. provided additional supervision and critical revisions.

The manuscript is a transparent account of the study being reported and adheres to PRISMA reporting guidelines. All listed authors have approved for the manuscript to be published in its current format and meet all the ICMJE criteria for authorship. The authors agree to be accountable for the contents of the paper and are jointly responsible for ensuring that all queries related to the accuracy or integrity of the project are investigated and resolved.

## ACKNOWLEDGEMENTS

We are grateful to Margot Kuylen for her contributions to the reliability assessment and to John Brazier for advising on the SF-36 to EQ-5D mapping function.

## ETHICAL APPROVAL

Not required.

## DATA SHARING

We have endeavoured to provide a detailed overview of the included studies and our analysis. Raw data is available upon request to the corresponding author.

